# Family Experiences, Challenges, and Coping Strategies During Women’s Perimenopause: A Qualitative Systematic Review Protocol

**DOI:** 10.1101/2025.09.21.25336300

**Authors:** Ravi Shankar, Jaminah Ali, Xu Qian

**Author notes:** **Corresponding Author:** Dr Ravi Shankar; Research and Innovation, Medical Affairs, Alexandra Hospital, Singapore, Email correspondence.

## Abstract

**Background:** Perimenopause represents a critical biopsychosocial transition affecting women globally, with profound impacts extending beyond individual experiences to encompass entire family systems. While research has documented various perimenopausal symptoms including vasomotor, psychological, and cognitive changes, limited attention has been given to how families as units experience and navigate this transition.

**Objectives:** This systematic review protocol aims to synthesize qualitative evidence on family experiences, challenges, and coping strategies during women’s perimenopause, addressing gaps in family-centered perspectives within existing literature.

**Methods:** Following PRISMA-P guidelines, we will search eight databases (PubMed, Web of Science, Embase, CINAHL, MEDLINE, The Cochrane Library, PsycINFO, and Scopus) from inception to October 2025. The PICo framework guides eligibility criteria focusing on family members of perimenopausal women, their experiences and coping strategies across diverse contexts. Covidence will facilitate study screening and selection. Quality assessment will employ the Critical Appraisal Skills Programme (CASP) checklist, with risk of bias evaluated using domain-based assessment. Thematic synthesis and meta-ethnographic approaches will analyze extracted data. CERQual will assess confidence in synthesized findings.

**Discussion:** This protocol anticipates generating comprehensive synthesis revealing family system impacts, typology of challenges, categorization of coping strategies, identification of cultural variations, and documentation of support system gaps. Evidence generated will inform development of family-centered interventions, clinical guidelines, and support systems addressing the multifaceted challenges families face during perimenopausal transitions.

## Introduction

Perimenopause represents a complex biopsychosocial transition affecting approximately one billion women globally, with cascading impacts on family systems that extend far beyond individual symptom experiences (1). This transitional phase, typically occurring between ages 40 and 58 and spanning several years before menstruation cessation, involves profound hormonal fluctuations manifesting as vasomotor symptoms, mood changes, sleep disturbances, and cognitive alterations that significantly influence family dynamics and relationships (2). Research indicates that perimenopausal women experience hot flashes, night sweats, irregular periods, and psychological symptoms that can profoundly affect their quality of life and interpersonal relationships (3). Despite affecting millions of families worldwide, research has predominantly focused on individual women’s experiences, overlooking the systemic family impacts of this universal life transition.

The intersection of perimenopausal changes with midlife stressors creates unique challenges for family systems. Women during this transition often manage multiple responsibilities including career demands, adolescent or young adult children, aging parents, and evolving marital relationships, creating what researchers term a “perfect storm” of midlife challenges (4). Family system theory posits that changes in one member inevitably affect all members, positioning perimenopause as a family developmental transition requiring systemic understanding and intervention (5). Partners frequently struggle to understand and respond supportively to the physical and emotional changes their spouse experiences, while children navigate their mother’s mood variability and changed energy levels during their own developmental transitions (6). Recent qualitative research has highlighted that women’s experiences of relationships during perimenopause reveal complex dynamics, with some reporting that “Friends? Supported. Partner? Not so much,” indicating differential support across relationship types (7).

Cultural and socioeconomic factors significantly shape how families experience and respond to perimenopause. Research demonstrates that women from different ethnic and cultural backgrounds experience varying symptom patterns, possess different explanatory models for understanding perimenopause, and utilize diverse coping strategies rooted in their cultural traditions (8). Studies have shown that “women from different ethnic groups may have different experiences from their White peers, such as entering the perimenopause and/or menopause at earlier ages” and having different symptom emphases (9). These cultural variations extend to family responses and support patterns, with some cultures viewing menopause as a natural transition deserving respect and support, while others may stigmatize or minimize the experience. Healthcare approaches have traditionally emphasized individual symptom management through hormone replacement therapy and other medical interventions, neglecting the relational dimensions requiring family-inclusive care models (10). The economic implications of perimenopause, including reduced work productivity and potential early workforce exit, create additional family stressors particularly when women are primary breadwinners (11).

### Objectives

This systematic review aims to synthesize qualitative evidence on family experiences, challenges, and coping strategies during women’s perimenopause, addressing critical gaps in understanding how families as systems navigate this transition. Primary objectives include identifying and characterizing experiences reported by different family members including partners, children, and extended family during perimenopausal transition; mapping the spectrum of challenges encountered by families navigating physical, psychological, and social changes associated with perimenopause; synthesizing evidence on coping strategies and support mechanisms utilized by families; exploring how cultural, ethnic, and socioeconomic factors influence family experiences and responses; and identifying gaps in existing qualitative literature to inform future research and practice development.

The review addresses several research questions examining family member experiences during women’s perimenopause and their impact on family dynamics and relationships. The primary research question asks: What are the experiences of family members during women’s perimenopause, and how do these experiences shape family dynamics and relationships? Secondary questions explore: What challenges do different family members face in understanding and responding to perimenopausal symptoms and changes? How do families cope with the physical, psychological, and social impacts of perimenopause? What support needs do families identify during the perimenopausal transition, and how are these needs currently being met or unmet? How do cultural, ethnic, and socioeconomic factors influence family experiences and coping strategies during perimenopause? What role do healthcare providers and systems play in supporting families during perimenopause? These questions align with recent research indicating that “knowledge, financial support, and family understanding are important to help women manage menopause” (12).

## Methods

### Study Design and Protocol Registration

This qualitative systematic review protocol follows the Preferred Reporting Items for Systematic Reviews and Meta-Analyses Protocols (PRISMA-P) 2015 guidelines ensuring comprehensive transparent reporting of planned methods (13). The protocol will be registered with the International Prospective Register of Systematic Reviews (PROSPERO) to promote transparency, reduce duplication, and minimize reporting bias. The review will employ the Enhancing Transparency in Reporting the Synthesis of Qualitative Research (ENTREQ) statement for reporting qualitative synthesis (14) and the Joanna Briggs Institute (JBI) methodology for systematic reviews of qualitative evidence (15). Any protocol amendments during the review process will be documented including rationales and potential impacts on findings, maintaining transparency throughout the research process.

### Conceptual Framework

The PICo framework (Population, phenomenon of Interest, Context) structures this systematic review, providing a comprehensive approach specifically designed for qualitative research questions (16). This framework guides all aspects of the review from eligibility criteria through data synthesis. The Population component encompasses family members of perimenopausal women including partners, children, parents, siblings, and other relatives, ensuring inclusive capture of diverse family perspectives rather than focusing solely on dyadic relationships. The phenomenon of Interest captures the multifaceted nature of family experiences during perimenopause, including emotional responses, communication patterns, coping strategies, support provision and receipt, and adaptation processes. The Context component acknowledges that family experiences of perimenopause occur across diverse settings including home environments, healthcare facilities, workplaces, and community spaces, while also considering cultural, socioeconomic, and geographical contexts that shape these experiences.

The PICo framework’s strength for this review lies in its flexibility and comprehensiveness for qualitative synthesis. Unlike frameworks designed for quantitative intervention studies, PICo accommodates the exploratory nature of qualitative research, allowing for emergence of unexpected themes while maintaining systematic rigor. The framework enables clear articulation of inclusion and exclusion criteria while remaining sensitive to the complexity and nuance inherent in family experiences. Its three-component structure ensures that the review captures not just who experiences perimenopause within families and what they experience, but crucially, the contextual factors that shape these experiences. This contextual sensitivity is particularly important given the cultural variations in how menopause is understood and experienced globally, and how family structures and dynamics vary across different societies.

### Eligibility Criteria

The population encompasses family members of women experiencing perimenopause, including partners of any gender, adolescent or adult children, parents, siblings, and other significant family members as defined by participants themselves. Studies focusing exclusively on perimenopausal women without inclusion of family member perspectives will be excluded, as the review specifically aims to capture family experiences rather than individual experiences alone. The phenomenon of interest includes family experiences, challenges, and coping strategies during perimenopause, encompassing family members’ understanding and interpretation of perimenopausal symptoms and changes; emotional, psychological, and relational impacts on family members; communication patterns and dynamics within families; support provision and receipt within family systems; coping strategies employed by families; and help-seeking behaviors and resource utilization. Studies that focus solely on clinical or medical aspects of perimenopause without addressing family experiences will be excluded from the review.

The context for this review includes all settings where families experience and manage perimenopause, including home environments, healthcare settings, workplace contexts, and community settings across all geographical locations and cultural contexts. Study designs comprise all qualitative methodologies including phenomenology, grounded theory, ethnography, narrative inquiry, case studies, participatory action research, and feminist research approaches. Mixed-methods studies will be included if the qualitative component can be extracted and meets inclusion criteria. Qualitative descriptive approaches without specified methodological frameworks providing rich descriptive data about family experiences will also be included. Quantitative studies, systematic reviews, opinion pieces, and clinical guidelines without primary qualitative data will be excluded. The review will include studies published from database inception to October 2025, capturing the full historical development of qualitative research on family experiences of perimenopause while including contemporary studies reflecting current family structures and dynamics.

**Table 1:**
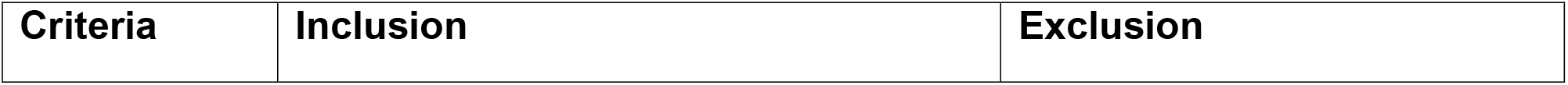
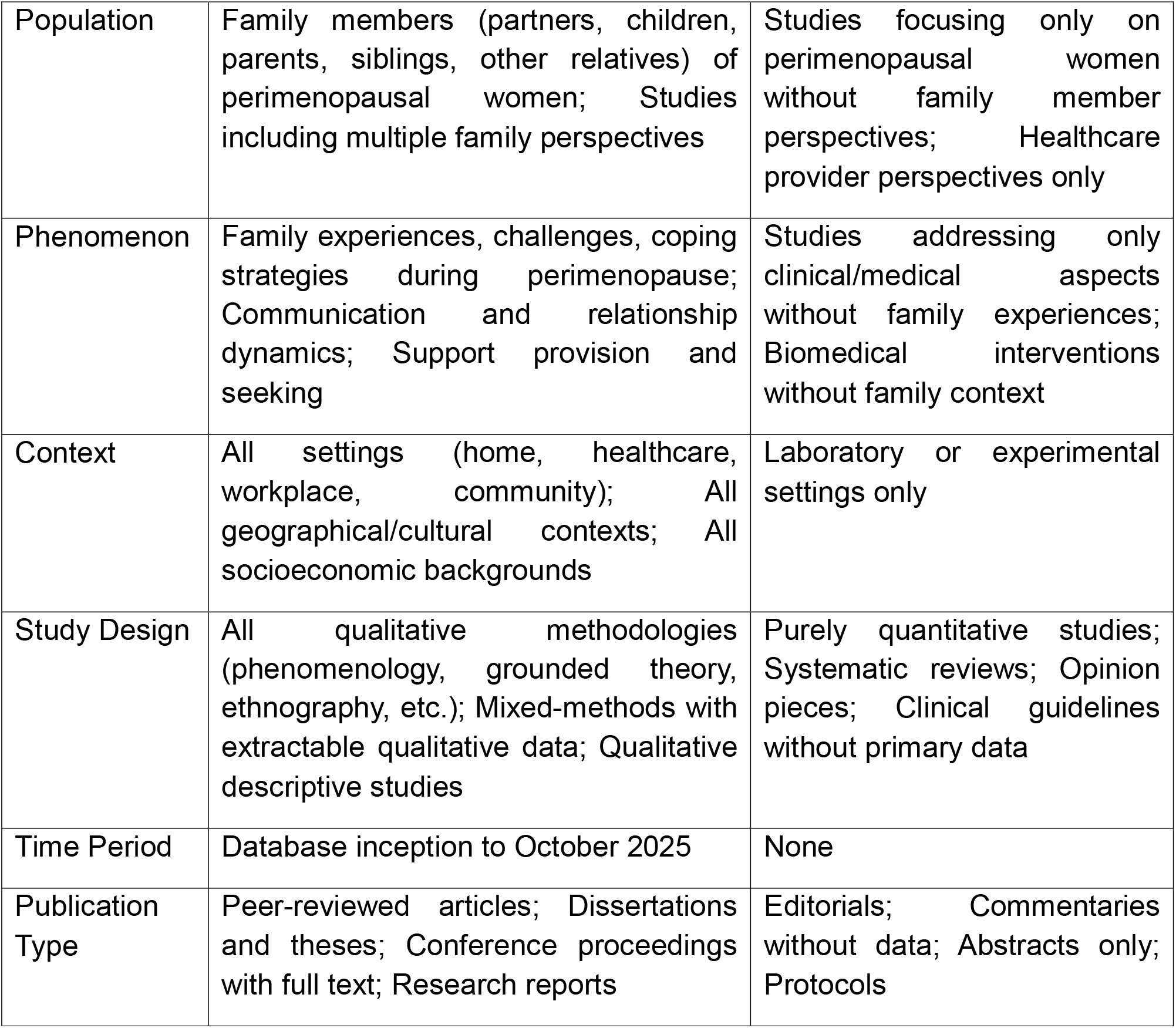
Inclusion and Exclusion Criteria.

### Information Sources and Search Strategy

Comprehensive database searching will encompass eight major health and social science databases to ensure interdisciplinary coverage: PubMed, Web of Science, Embase, CINAHL, MEDLINE, The Cochrane Library, PsycINFO, and Scopus, searched from inception to October 2025. This temporal scope aligns with recommendations for comprehensive systematic reviews to capture the full evolution of qualitative research on the topic (17). Grey literature sources will include ProQuest Dissertations and Theses Global, OpenGrey, Conference Proceedings Citation Index, and institutional repositories of universities known for menopause and family research. Supplementary strategies involve forward and backward citation searching of included studies, reference list hand-searching, expert consultation with researchers in menopause and family studies, and monitoring research networks including ResearchGate and Academia.edu for relevant studies.

The search strategy employs a three-concept approach adapted from established systematic review methodologies: ((‘perimenopause’ OR ‘menopaus^*^ transition’ OR ‘climacteric’ OR ‘midlife transition’ OR ‘menopausal symptoms’ OR ‘hot flash^*^‘ OR ‘hot flush^*^‘ OR ‘night sweat^*^‘ OR ‘hormone^*^ chang^*^‘ OR ‘estrogen declin^*^‘ OR ‘reproductive aging’) AND (‘family’ OR ‘partner^*^‘ OR ‘spouse^*^‘ OR ‘husband^*^‘ OR ‘wife’ OR ‘wives’ OR ‘children’ OR ‘offspring’ OR ‘son^*^‘ OR ‘daughter^*^‘ OR ‘parent^*^‘ OR ‘mother^*^‘ OR ‘father^*^‘ OR ‘sibling^*^‘ OR ‘relationship^*^‘ OR ‘marital’ OR ‘interpersonal’ OR ‘family support’ OR ‘family experience^*^‘ OR ‘family challenge^*^‘ OR ‘family coping’ OR ‘family adaptation’ OR ‘family dynamics’ OR ‘family functioning’ OR ‘caregiv^*^‘ OR ‘social support’ OR ‘emotional support’) AND (‘qualitative’ OR ‘interview^*^‘ OR ‘focus group^*^‘ OR ‘thematic analysis’ OR ‘phenomenolog^*^‘ OR ‘grounded theory’ OR ‘ethnograph^*^‘ OR ‘content analysis’ OR ‘narrative^*^‘ OR ‘lived experience^*^‘ OR ‘discourse analysis’ OR ‘interpretive’ OR ‘constructivist’ OR ‘hermeneutic^*^‘)). Search strategies will be adapted for each database while maintaining conceptual consistency, with assistance from a health sciences librarian experienced in systematic review searching.

### Study Selection

Covidence systematic review software will manage the entire selection process, providing systematic tracking and documentation of all screening decisions (18). Two independent reviewers will conduct title and abstract screening followed by full-text assessment using standardized screening forms. Calibration exercises using 50 randomly selected citations will ensure consistent application of eligibility criteria, with reviewers discussing discrepancies to achieve consensus before proceeding with full screening. Disagreements during screening will be resolved through discussion, with third reviewer consultation when consensus cannot be reached. Inter-rater reliability will be assessed using Cohen’s kappa statistics, targeting a minimum agreement of 0.75 indicating substantial agreement (19).

Full-text screening will involve detailed assessment against all eligibility criteria, with specific reasons for exclusion documented for transparency. Authors of studies will be contacted when additional information is needed to determine eligibility, with a maximum of three contact attempts over a four-week period before excluding studies due to insufficient information. A PRISMA flow diagram will document the complete selection process, including number of citations identified from each source, citations excluded at each stage, and primary reasons for exclusion at full-text screening, ensuring transparency and reproducibility of the selection process (17).

### Data Extraction

Standardized data extraction forms developed specifically for this review will capture comprehensive information about study characteristics, methodological approaches, participant demographics, and findings relevant to family experiences of perimenopause. The extraction form will be piloted on five randomly selected included studies and refined based on this pilot before full extraction begins. Two reviewers will independently extract data from each included study, with discrepancies resolved through discussion and verification against original studies. Study characteristics to be extracted include authors, year of publication, country and setting, study aims and research questions, theoretical or conceptual frameworks employed, funding sources, and any declared conflicts of interest.

Methodological information will encompass the qualitative methodology used (phenomenology, grounded theory, ethnography, etc.), sampling strategy and sample size, data collection methods (individual interviews, focus groups, observation, document analysis), data analysis approach, strategies used to enhance trustworthiness or rigor, and reflexivity statements. Participant characteristics will be documented in detail, including number and types of family members included, demographic information (age, gender, ethnicity, education, socioeconomic status), relationship to the perimenopausal woman, duration of the perimenopause experience, and any relevant contextual factors such as living arrangements or caregiving responsibilities.

Findings extraction will focus on themes, concepts, and insights related to family experiences, challenges, and coping strategies during perimenopause, with particular attention to direct participant quotations that illustrate key findings. First-order constructs (participant voices) will be distinguished from second-order constructs (author interpretations) to maintain analytical clarity (20). Contextual factors influencing family experiences, including cultural beliefs, social norms, economic circumstances, and healthcare system factors, will be carefully documented to enable exploration of how context shapes family responses to perimenopause.

### Quality Assessment

The Critical Appraisal Skills Programme (CASP) Qualitative Research Checklist will assess study quality, examining ten key domains: clarity of research aims, appropriateness of qualitative methodology, research design suitability, recruitment strategy appropriateness, data collection methods, researcher-participant relationship consideration, ethical considerations, rigor of data analysis, clarity of findings presentation, and overall research value (21). Two reviewers will independently assess quality with disagreements resolved through discussion and third reviewer consultation when necessary. Rather than using quality scores for study exclusion, assessment will inform sensitivity analyses exploring whether findings differ based on study quality levels.

Additional quality considerations specific to family research will include assessment of whether multiple family member perspectives were included and how they were integrated; how family dynamics were captured and analyzed beyond individual experiences; whether researchers considered the family as a system rather than collection of individuals; how power dynamics within families were addressed; and attention to reflexivity regarding researchers’ own experiences with perimenopause or family relationships. Studies will be categorized as high quality (meeting 8-10 criteria), moderate quality (meeting 5-7 criteria), or low quality (meeting fewer than 5 criteria), with particular weight given to analytical rigor and depth of family perspective exploration.

### Risk of Bias Assessment

Risk of bias will be evaluated using a domain-based assessment approach adapted for qualitative research (22). Selection bias will be examined through recruitment strategies assessment, examining whether participants represent diverse family experiences or reflect limited perspectives, and whether recruitment methods might have systematically excluded certain family types. Detection bias assessment will consider data collection methods appropriateness for capturing family experiences, researcher reflexivity and potential influence on data generation, and whether data collection allowed all family members to express their perspectives freely. Attrition bias will evaluate participant retention throughout studies, completeness of data from all recruited participants, and whether missing data might systematically affect certain perspectives.

Reporting bias will be assessed through examination of selective outcome reporting, transparency in reporting both confirmatory and contradictory findings, and completeness of demographic and contextual information reporting. Other potential biases include funding source influence, researcher conflicts of interest, and temporal bias related to when studies were conducted. Each domain will be rated as low, moderate, or high risk with detailed justifications documented. Overall bias risk will be determined by considering all domains collectively, with studies rated as low risk if most domains show low risk, moderate risk if domains show mixed risk levels, and high risk if multiple domains show high risk. Sensitivity analyses will explore whether synthesis findings differ when high-risk studies are excluded.

### Data Synthesis

Thematic synthesis following Thomas and Harden’s three-stage approach will be complemented by meta-ethnographic techniques for translating concepts across studies (20, 23). The first stage involves line-by-line coding of findings sections including participant quotations and author interpretations, with initial coding remaining close to the data using participants’ own terms where possible. Two reviewers will independently code a subset of five studies, comparing coding to develop an initial framework that will be applied to all studies with new codes added as necessary. NVivo qualitative data analysis software will facilitate coding management and team collaboration throughout the synthesis process.

The second stage involves organizing codes into descriptive themes capturing patterns across studies through constant comparison to identify similarities and differences in family experiences, challenges, and coping strategies. Attention to deviant cases and contradictory findings will explore factors accounting for variation in family experiences. Descriptive themes will remain close to original study findings, providing comprehensive mapping of existing knowledge about family experiences of perimenopause. The third stage develops analytical themes going beyond individual study findings to generate new interpretive insights, drawing on meta-ethnographic techniques for concept translation across studies and synthesis of translations to develop third-order constructs.

Contextual factors including cultural background, socioeconomic status, healthcare system characteristics, and geographical location will be examined through subgroup analyses exploring whether family experiences and coping strategies differ across contexts. Temporal patterns throughout the perimenopausal transition will be analyzed to identify critical periods or turning points in family adaptation. The synthesis will maintain an audit trail documenting all analytical decisions and interpretations to ensure transparency and enable assessment of synthesis trustworthiness.

### Confidence Assessment

The Confidence in Evidence from Reviews of Qualitative research (CERQual) approach will assess confidence in synthesized findings based on four components (24). Methodological limitations will be assessed by examining the extent to which studies contributing to a finding have methodological problems based on CASP assessments. Coherence evaluation will determine whether findings are well-grounded in data from contributing studies and provide convincing explanations for patterns found across studies. Data adequacy assessment will examine whether studies provide sufficiently rich data and whether the body of evidence is thin or limited in quantity. Relevance appraisal will evaluate the extent to which evidence from contributing studies is applicable to the review question’s context, population, and phenomenon of interest.

Each synthesized finding will receive an overall confidence rating of high (minor concerns across components), moderate (moderate concerns in one or more components), low (substantial concerns in one or more components), or very low (serious concerns across multiple components). Two reviewers will independently conduct CERQual assessments with disagreements resolved through discussion. Summary of Qualitative Findings tables will present each finding with its CERQual assessment and detailed explanation of the judgment, enabling users to understand evidence strength supporting each finding and make informed decisions about applicability to their contexts.

## Discussion

This protocol addresses critical gaps in perimenopause literature by adopting family systems perspectives previously overlooked in research focusing predominantly on individual women’s experiences. The comprehensive methodology ensures rigorous evidence synthesis while maintaining sensitivity to diverse family experiences across cultural contexts. Recent research has emphasized that “the menopausal experience extends beyond physical symptoms, also affecting mental health, personal and professional life, and self-identity” with clear implications for family systems (25). By centering family perspectives, this protocol challenges prevailing biomedical paradigms treating perimenopause as individual medical events rather than family developmental transitions.

The PICo framework provides robust structure for this systematic review, ensuring comprehensive capture of complex family dynamics during perimenopausal transition. Unlike frameworks designed for quantitative intervention studies, PICo’s qualitative focus aligns perfectly with the review’s aims to understand lived experiences, meanings, and contextual influences on family adaptation. The framework ensures appropriate study identification while maintaining flexibility for diverse qualitative methodologies, from phenomenological explorations of lived experience to ethnographic examinations of cultural influences on family responses. This framework-guided approach represents methodological rigor in qualitative synthesis, providing clear structure while preserving the interpretive depth essential for understanding family experiences of perimenopause.

The protocol’s strength lies in its inclusive approach encompassing diverse family member perspectives rather than privileging particular viewpoints. By searching databases from inception to October 2025, the review captures historical evolution of family perimenopause research while including contemporary studies reflecting current family structures and societal attitudes. This temporal scope is particularly important given rapidly evolving social perspectives on menopause and increasing recognition of its impacts beyond individual women. Covidence utilization ensures systematic transparent study selection minimizing bias and enhancing reproducibility, addressing common criticisms of qualitative synthesis regarding selection transparency (26).

The comprehensive search strategy including grey literature reduces publication bias risk while capturing diverse research contexts often excluded from traditional database searches. This is particularly important for family perimenopause research which may appear in disciplines beyond health sciences, including sociology, anthropology, and family studies. The adapted search string’s three-concept approach balances sensitivity and specificity, ensuring relevant study capture without overwhelming reviewers with irrelevant citations.

Quality assessment using CASP alongside risk of bias evaluation provides nuanced understanding of evidence strengths and limitations. Rather than excluding lower-quality studies potentially containing valuable insights, sensitivity analyses will explore quality impacts on findings, maximizing evidence utilization while maintaining analytical rigor. This approach acknowledges qualitative research’s interpretive nature while ensuring synthesis credibility. The additional assessment criteria specific to family research ensure that studies adequately capturing family dynamics receive appropriate weight in the synthesis.

Several limitations warrant acknowledgment. The synthesis quality depends on primary study quality and comprehensiveness; if studies inadequately capture family perspectives or focus narrowly on specific family members, synthesis will be similarly limited. The review may be affected by historical lack of attention to family experiences in perimenopause research, potentially resulting in smaller eligible study pools than topics receiving more research attention.

The protocol’s emphasis on family-centered perspectives has important implications for healthcare delivery, potentially transforming service design to incorporate family needs and perspectives. Evidence suggests that “healthcare providers may not be trained to recognize perimenopausal and post-menopausal symptoms and counsel patients on treatment options” and even less prepared to address family dimensions. Findings may support development of family-inclusive clinical guidelines, educational programs for family members, and training for healthcare providers in family-centered menopause care.

Workplace implications include supporting arguments for flexible policies accommodating not just perimenopausal women but family members providing support. Research indicates that “severity of the physical symptoms of perimenopause affect job performance, and severity of psychological symptoms affect job retention” with clear implications for family economic stability (27). Understanding family impacts strengthens cases for comprehensive employee assistance programs and workplace education promoting understanding among colleagues and supervisors.

Policy implications extend to healthcare system design, insurance coverage for family support services, and community program development recognizing perimenopause as family transitions requiring systemic support. The synthesis may reveal successful international models for family-centered perimenopause support that could inform policy development in other contexts. Evidence of family impact may strengthen advocacy for increased research funding and service development in this neglected area.

This protocol contributes to theoretical understanding of family adaptation to health transitions, potentially extending existing resilience and coping theories. The meta-ethnographic approach may generate new theoretical insights about how families navigate midlife health transitions, contributing to broader family development and adaptation literature. Methodological contributions include demonstrating multi-framework application in qualitative synthesis and comprehensive bias assessment in family-focused health research, providing template for future systematic reviews in similar domains.

Ethical considerations permeate the protocol despite involving published research synthesis rather than primary data collection. Maintaining participant dignity from original studies requires careful contextualized use of quotations avoiding misrepresentation or stigmatization. Equity attention ensures diverse population representation with explicit acknowledgment when groups are underrepresented. Team reflexivity regarding personal perimenopause experiences ensures data-grounded synthesis rather than preconception-based interpretations, particularly important given the gendered nature of perimenopause and potential for researcher bias.

Future implications include identifying research priorities addressing evidence gaps, particularly regarding underrepresented family member perspectives such as adolescent children, male partners, and extended family members. Longitudinal research examining how family experiences evolve throughout perimenopausal transition represents another priority area. Methodological insights will improve future qualitative research design and conduct in family health contexts, potentially leading to more rigorous and comprehensive studies. The protocol establishes foundation for generating evidence transforming understanding and support for families navigating perimenopause, ultimately improving quality of life for perimenopausal women and their families through comprehensive understanding of their lived experiences, challenges, and strengths.

## Data Availability

All data produced in the present work are contained in the manuscript

